# Visual preferences for communicating modelling: a global analysis of COVID-19 policy and decision makers

**DOI:** 10.1101/2024.11.05.24316774

**Authors:** Liza Hadley, Caylyn Rich, Alex Tasker, Olivier Restif, Sebastian Funk

## Abstract

Effective communication of modelling results to policy and decision makers has been a longstanding challenge in times of crises. This communication takes many forms - visualisations, reports, presentations - and requires careful consideration to ensure accurate maintenance of the key scientific messages. Science-to-policy communication is further exacerbated when presenting fundamentally uncertain forms of science such as infectious disease modelling and other types of modelled evidence, something which has been understudied. Here we assess the communication and visualisation of modelling results to national COVID-19 policy and decision makers in 13 different countries. We present a synthesis of recommendations on what aspects of visuals, graphs, and plots policymakers found to be most helpful in their COVID-19 response work. This work serves as a first evidence base for developing guidelines on the communication and translation of modelling-to-policy.

## 1 Introduction

Effective communication of scientific results to policy and decision makers has been a longstanding challenge in times of crises [1,2]. Both the language and timelines of science and policy are fundamentally distinct, leading to tensions and difficulties in a cohesive working relationship. This communication is further challenged in the field of epidemiology and modelling (referred to henceforth as ‘modelling’ for brevity), where results are by definition uncertain and must be interpreted carefully. Here we assess the communication and visualisation of modelling results to national COVID-19 policy and decision makers in 13 different countries. This work serves as a first evidence base for developing guidance on communicating modelling and broader scientific results to policymakers.

**Figure 1:**
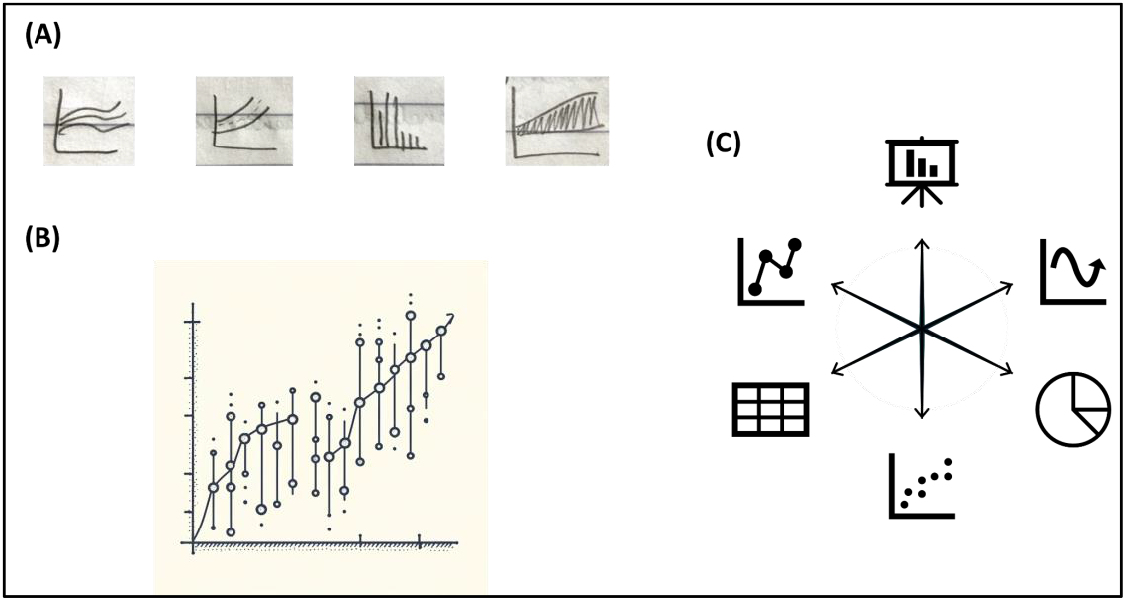
There are many decisions to make when designing visuals for policy communication. Depicted are: (A) hand-drawn sketches, (B) an AI-generated graph displaying uncertainty, created with DALL·E 3, (C) graphic icons, used with permission from Microsoft.

Looking back to the 2009 Influenza pandemic, mathematical modellers and biostatisticians involved were found to provide advice to their countries mainly through advisory groups consisting of multiple disciplines [3]. Van Kerkhove & Ferguson commented that while sophisticated modelling analyses were carried out regularly, it was actually the simpler real-time statistical analyses using available epidemiological and virological data that were often most helpful in informing daily policy decisions. The authors also discuss communication on the science-policy interface and described communication between modelling groups and policy-makers in 2009 as “good in several countries but could be improved further”. More recent works have looked at communication with the public in crises [4–6] but while there is a plethora of reporting on generic data visualisations in COVID-19 [7], research that examines modelling visualisations and the translation of epidemiological modelling results *as understood by policymakers* is more sparse.

One study on this theme is that of the inter-agency COVID-19 Multi-model Comparison Collaboration (CMCC) [8,9]. Their work highlights the difficulties of communicating and working with modelled evidence, and introduces practical suggestions for effectively reporting and communicating model results taken from a rapid stakeholder survey carried out in 2020. Additional research into visualisation, communication, and framing of modelling is now required. In particular, there is scope to substantiate the work of the CMCC through in-depth qualitative interviews in a wide range of countries, teasing out choice of language and preferred metrics when communicating modelling and identifying aspects of plots and graphs that are (reported to be) well-understood or not well-understood by policy and decision makers.

The aim of this study is hence to identify policymakers’ visual preferences and advice for translating modelling *to policy* in times of crises, expanding on the initial CMCC work highlighted above. We report below a qualitative analysis of findings using thematic analysis [10]. For results considering the wider interface of epidemiological modelling and policymaking, and recommendations for future practices, see adjacent work [11]. The focus of COVID-19 in this study provides a concrete example of wide-reaching scientific influence in policy spheres, so while some results have an epidemiological focus in places, the high-level findings will be widely motivating and applicable to other science-policy systems.

## 2 Methods

### 2.1 Study design and recruitment

We carried out a qualitative interviews study to understand the interaction of COVID-19 mathematical modelling and policymaking in different countries. Methods are described in full-detail in [11] and are summarised here. In total, 12 countries and 1 jurisdiction (Hong Kong, China) took part in the study.^1^ Target countries were chosen through stratified random sampling by UN geographic region to select a first country in each region, with subsequent countries chosen to capture the breadth of COVID-19 experience including differing levels of: income status, pre-COVID-19 pandemic preparedness ratings, population size, COVID-19 prevalence, prior experience of epidemiological modelling for national decision-making (including none), and modelling infrastructures or consortiums (including those with no formal setup). Countries with no reported epidemiological modelling activity informing their national decision making at any level during COVID-19 were excluded from the study as they would not meet our eligibility criteria (see Hadley et al., 2024 Supplementary Material for further details). Countries with a population of less than five million were also excluded. The study researchers then identified suitable interviewees in each target country via purposeful sampling of key modelling-to-policy actors, recruiting epidemiological modellers and the national science advisors and policy/decision makers (i.e. model users) that these modellers interacted with [12]. If no relevant interviewees could be sought, a second country was chosen instead. Overall, 27 interviewees from 13 countries took part. Twelve of these individuals self-identified as scientific advisors or policy/ decision makers (3, 9 respectively), and 15 as epidemiological modellers. Details of the study countries and make-up of study participants are shown in Table 1 and Table 2 respectively.

**Table 1:** The 13 countries and jurisdictions involved in our study and relevant demographic statistics. Each interviewee reported their country(ies) of practice as one of the above. LAC = Latin America and the Caribbean; LIC = Low-income country; LMIC = Lower-middle income country; MIC = Upper-middle income country; HIC = High-income country.

| Country of practice | Country details |  |  |  |
| --- | --- | --- | --- | --- |
|  | UN geographic region<sup>2</sup> | Income status<sup>3</sup> | GHS pandemic preparedness rating (2019)<sup>4</sup> | Population size (in millions)<sup>5</sup> |
| Australia | Oceania | HIC | 4th | 25.8 |
| Canada | Northern America | HIC | 5th | 38.0 |
| Colombia | LAC | UMIC | 65th | 51.2 |
| France | Europe | HIC | 11th | 64.5 |
| Hong Kong, China<sup>6</sup> | Asia | HIC | 51st* | 7.5 |
| Japan | Asia | HIC | 21st | 124.9 |
| Kenya | Africa | LMIC | 55th | 52.5 |
| New Zealand | Oceania | HIC | 35th | 5.1 |
| Peru | LAC | UMIC | 49th | 33.5 |
| South Korea | Asia | HIC | 9th | 51.8 |
| South Africa | Africa | UMIC | 34th | 59.1 |
| Uganda | Africa | LIC | 63rd | 45.1 |
| United Kingdom | Europe | HIC | 2nd | 67.2 |

<sup>2</sup> UN geographic region as defined in: United Nations World Population Prospects 2022 Locations Documentation, Online Edition [13].
<sup>3</sup> Income as defined by World Bank income groups [13].
<sup>4</sup> Ranking taken from the 2019 Global Health Security Index, indicating expected overall preparedness for an infectious disease outbreak [14]. All countries are above the average score ranking of 85th position. \*Note: Hong Kong's ranking of 51st represents the overall ranking for China.
<sup>5</sup> Population as of 1<sup>st</sup> January 2021, taken from United Nations World Population Prospects 2022 Demographic Indicators, Online Edition [15].
<sup>6</sup> As noted above, Hong Kong is a Special Administrative Region of China.

**Table 2:**
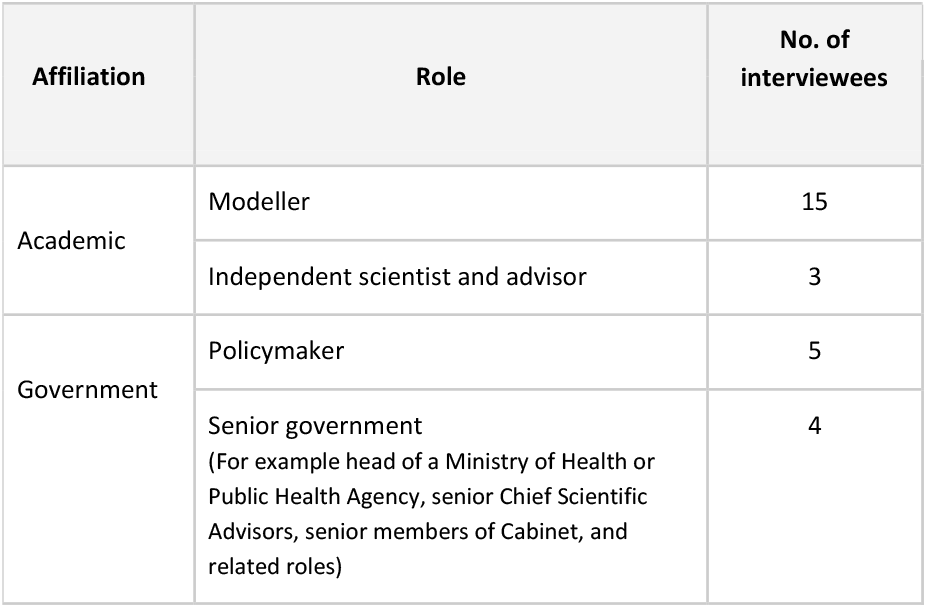
Make-up of study participants. The term ‘academic modeller’ refers to researchers in epidemiological modelling as noted in the main text.

### 2.2 Interview format

Interviews were in-depth semi-structured, lasting on average one hour in length. The majority were carried out by video conferencing software, with four being carried out in-person. In the interview, the interviewer facilitated structured discussion on four topics in turn: Structures and pathways to policy, Collaboration and knowledge transfer, Communication and visualisation, and Evaluation and reflection. See the Supplementary Material for an example interview schedule, which was motivated by earlier work of the research team [16]. This paper concentrates on the topic of communication and visualisation, while adjacent work [11] presents a fuller analysis of the interactions of modelling and policy in each country.

Interview prompts on the topic of communication and visualisation began with example-based questions such as “Can you give an example of a plot that you felt worked well for communicating model findings?” and “Can you give an example of a plot that was misunderstood or not quite so effective at demonstrating model findings?”. The interviewer then led interviewees into a wider discussion on the specific aspects of plots and graphs that policy and decision makers found most helpful for communicating epidemiological findings. Interviews also covered preferred language/metrics for graphics and a discussion on what modellers should include or exclude in reports and presentations to policy, such as the level of detail and how accurate one should be.

### 2.3 Data analysis

All interviews were audio recorded, transcribed, qualitatively coded, and reviewed for accuracy by the study team. Analysis of the full interview data followed thematic analysis as described in Braun & Clarke [10], and findings specifically related to visualisation were synthesised and presented here. A draft version of findings was also shared with interviewees prior to publication to allow for revisions and corrections. This research study has been reviewed by the University of Cambridge Psychology Research Ethics Committee (application number PRE.2023.034). All participants provided informed consent.

## 3 Results

When presenting scientific results to decision makers, one has several choices in terms of what to emphasise and how best to represent that information visually and verbally. Below we present a detailed summary of the visual preferences as identified from our respondents; content represents a synthesis of ideas from national policymakers, science advisors, and epidemiological modellers on what visual aspects were helpful for *policy* in COVID-19. We conclude with real-world examples from COVID-19 modelling teams (Figures 2-5).

**Figure 2:**
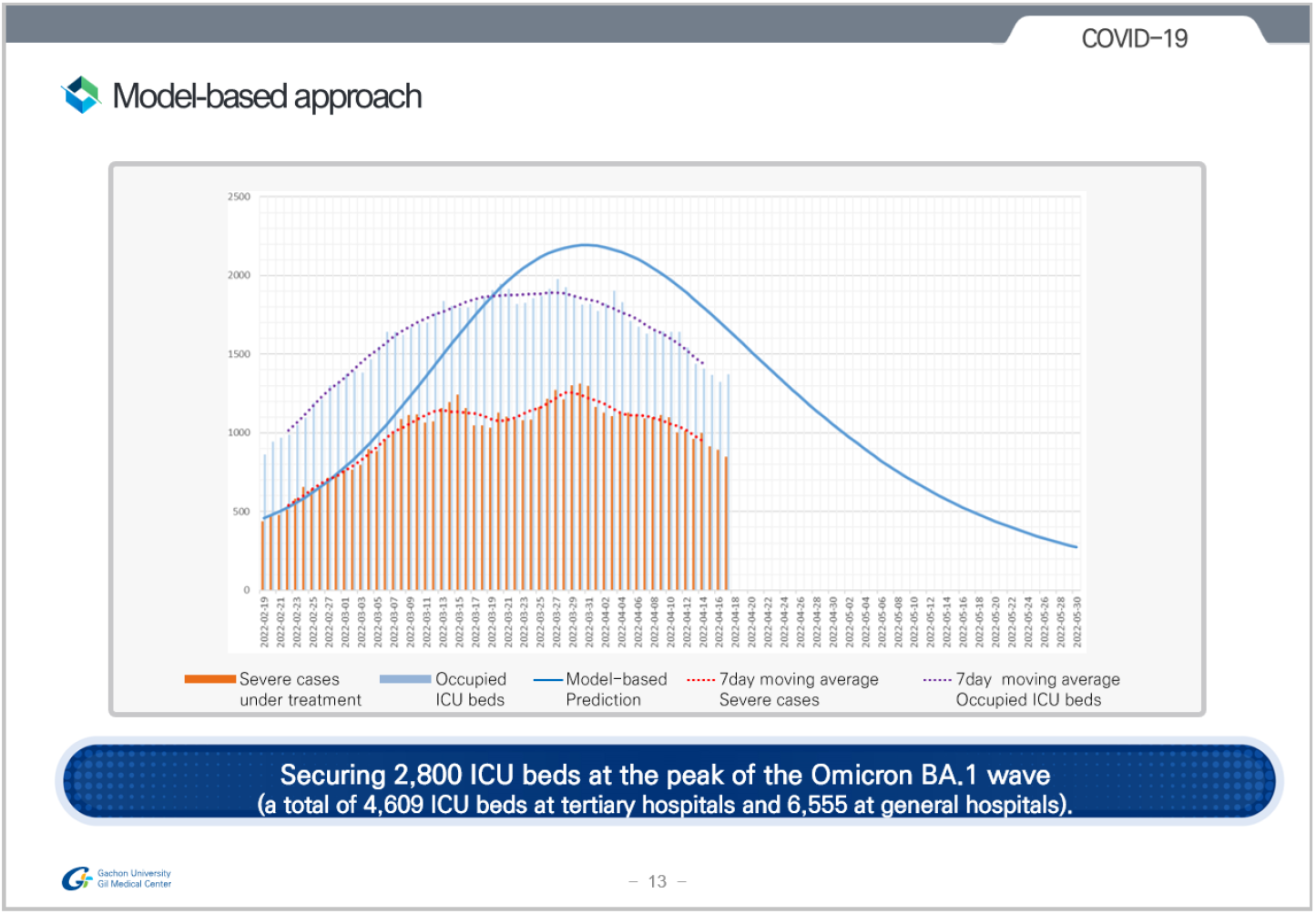
Example of the graph format used by South Korean modellers when presenting findings to decision makers. Slide depicts predictions on ICU bed numbers for the Omicron wave in April 2022. Reproduced with permission from the authors.

**Figure 3:**
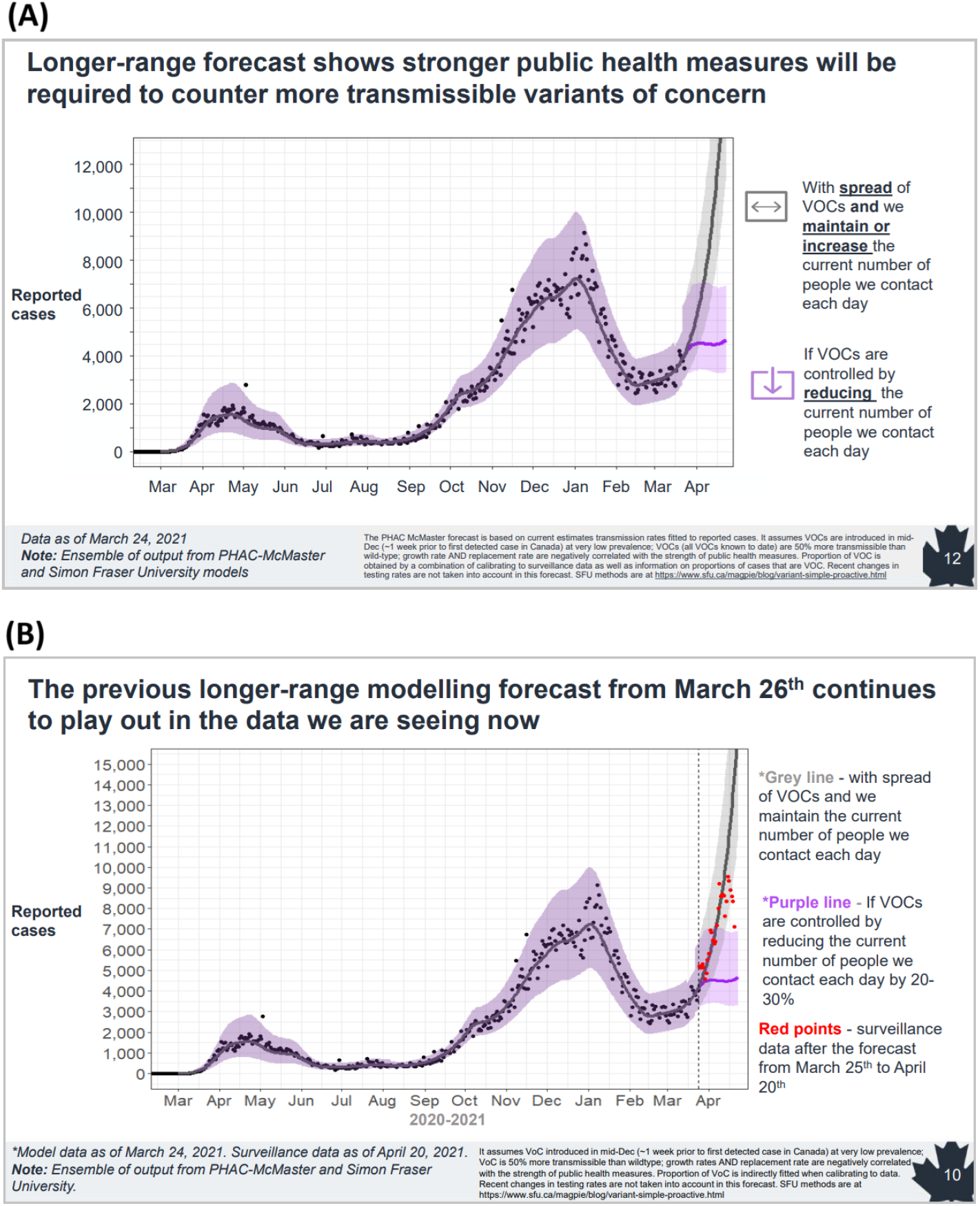
Examples of modelling visuals from Canada [17]. (A) Figure presented in one of the Epidemiology and Modelling COVID-19 Technical Briefings from the Public Health Agency of Canada in March 2021. Slide shows mathematical model-based long-range forecasting of the impact of the Alpha variant. The key message is displayed as a title. (B) The same visualisation one month on, in April 2021 with March data overlaid, again emphasising key findings in a clear manner. Reproduced with permission from the Public Health Agency of Canada.

**Figure 4:**
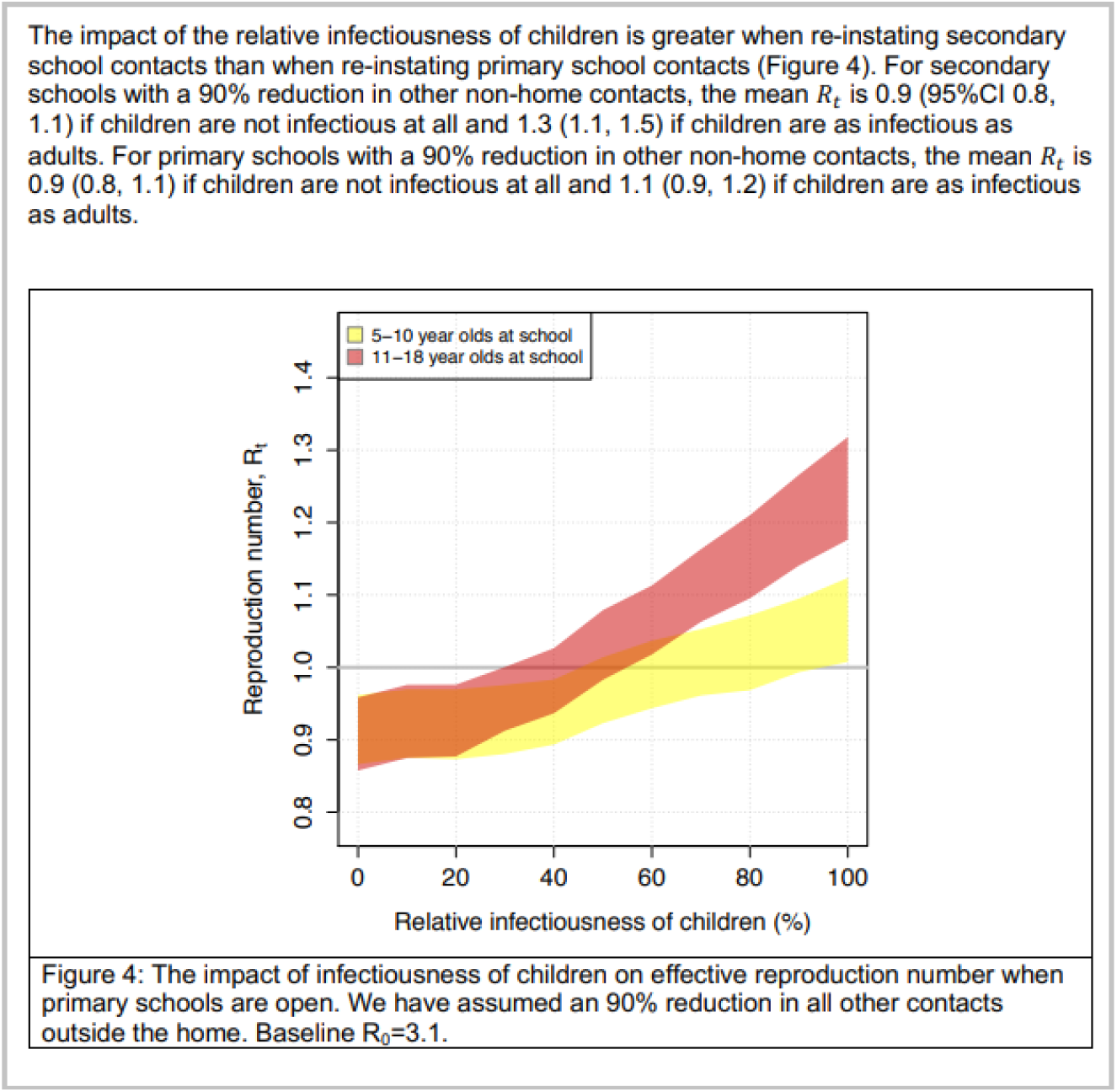
Example of a ‘watermelon slice’ diagram from members of the UK modelling consortium SPI-M-O. This figure is taken from an early modelling report of the University of Bristol and University of Exeter on the impact of opening schools in late April 2020 [18]. Figure compares opening primary schools with opening secondary schools, when the infectiousness of children was unknown. Figures such as these, accompanied by clear summary statements, helped illustrate key modelling findings in official SPI-M-O Consensus Statements to policy. Reproduced with permission.

**Figure 5:**
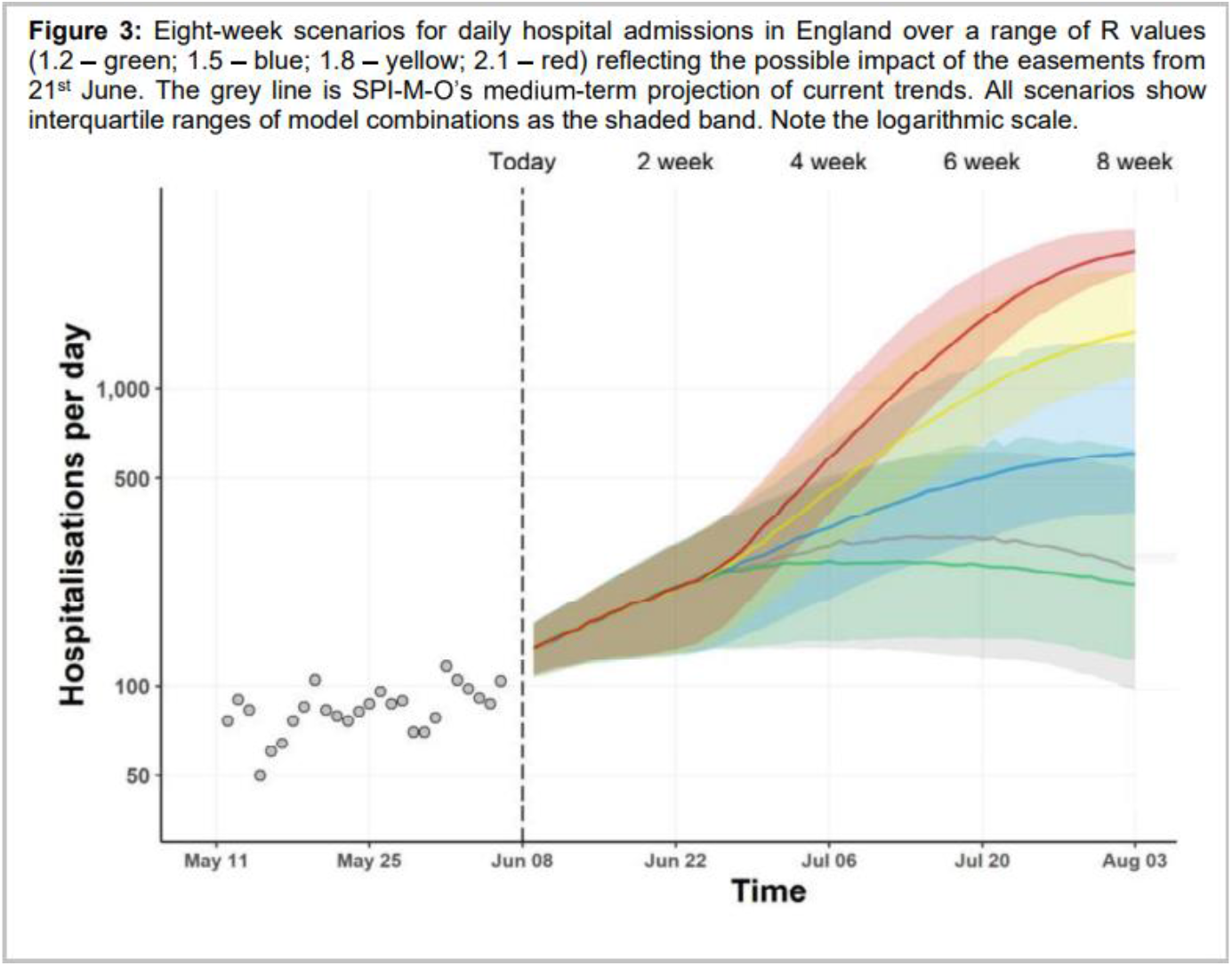
Example of a ‘rainbow diagram’ generated by the UK modelling consortium SPI-M-O. Figure shows extrapolations for different plausible levels of the effective reproduction number Rt after an easing of restrictions in the UK in mid 2021. The figure was accompanied by clear summary statements outlining the major modelling findings (not shown). Taken from SPI-M-O Consensus Statement 9th June 2021 [19]. Reproduced with permission from the SPI-M-O Secretariat.

### Overarching findings

#### Be cautious with over-persuasion

Visuals were deemed important in many settings, but it is worth first noting that there was a hesitancy to their use by a few individuals. One interviewee expressed concern that “models can be very persuasive to people who are not modellers”. With lots of wonderful coloured moving maps, “it is very hard for people not to believe they are true”. This was reciprocated by other respondents who noted that “graphs are always seen as a prediction by non-modellers”. Advice was given on interpreting and reading graphs to combat this (see ‘Graph-reading’).

#### Graphs with trends helpful; mixed opinions on ‘fancy graphics’

Some policymakers described specific types of graphs that were useful to them: graphs with trends, graphs with dates, tables and number-driven discourse, and trends and scenarios. Key visuals in the Office for the President in Uganda for example included bar graphs, pie charts, line graphs, and short video clips and animations. Interactive maps and dashboards were reported to be useful too. However there were mixed views in other countries - interactive graphics used early on in Kenya were well-received, but advisors in South Korea warned of their overuse: “Modellers sometimes strive for fancy graphics - heatmaps, dashboards, interactive elements and so on but I disagree. The most important visualisation in my experience is a simple time-series epidemiological curve and its future prediction curve”. This can avoid confusion in time-constrained settings.

#### Oversharing is useless

Several others agreed with the above line of thinking. As one respondent from Australia commented, “it is critical to understand that oversharing of information (i.e. scientific detail) is perceived as useless to policymakers”. Others echoed these points, stressing the distinction between what is academically important and what is important in policy. “Scientists need to recognise policy makers don’t have time/ capacity for long-winded answers!”. Being able to give a ballpark figure was very helpful. For example, another remarked that “providing confidence intervals through Bayesian approaches to future predictions or bootstrapping is academically important but not important in policy”. Intermediaries in the UK and South Africa acknowledged these challenges and portrayed the “difficult dance” of communication between science and policy, balancing the need for brevity with the need to understand the level of uncertainty and relevant limits of the scientific research each week. One piece of advice was to rule out the less useful parts of modelling; in a few countries presenting numbers of cases for example was quickly deemed ineffective, since this metric is not accurate and is heavily dependent on level of testing. Some modellers for example stopped presenting and communicating cases after their first workpackage.

#### Desire for actionable estimates

Decision makers in Hong Kong and mainland China remarked that they were confused by wide confidence intervals and frequently requested actionable estimates. Modellers were asked instead for the operational analysis, i.e interpretation of the estimates in terms of what it means for policy. Advisors in New Zealand also stressed the need for modellers who understood that diagrams with extremely wide confidence intervals were not helpful, dubbed “crayon diagrams” - diagrams with huge uncertainty that could have been drawn with a crayon.

#### Re-translation will occur

Several respondents also identified a focus on whether what modellers explained could be easily explained and translated again by non-modellers (i.e. by the intermediary, policymaker, or other relevant non-modelling expert advisor through layers of government). This is especially important in larger country settings with multi-faceted or multi-layered government systems. We introduce the term ‘re-translation’ to describe this activity and acknowledge the distinction between first translation from modeller to non-modeller, and subsequent re-translations where maintaining both message and nuance is crucial.

### Specific aspects of graphs

#### Simplicity is key

When asked what types of visuals were helpful to decision makers, the vast majority of interviewees without prompt first stressed the need for *simple graphics*. Modellers aimed to present epidemiological findings in simple terms, often a simple line graph epidemiological curve in the language or metrics that their respective decision makers were familiar with. Further discussion on ‘Metrics’ is in the subsequent section while specific aspects of graphs that were found to be helpful are described here.

#### Options were found to be helpful

‘Scenario modelling’ thinking was reportedly absorbed well by decision makers in many of the countries in the study. The interviewees from France, Kenya, Japan, South Korea, UK, and Colombia all reported that modellers presented ‘options’ or ‘scenarios’ to decision makers, often in the form of a simple repeated epidemic curve showing variation in a key parameter. E.g., “Let’s vary the reproduction number. If it’s this versus this versus this, this is what happens”. Figures that will give the policymaker options (i.e., data, calibration to the data, and then the different options/scenarios) were found to be most helpful, and there was a high demand for this type of scenarios work for planning and resource allocation (“If A, then B. If C, then D. How many beds? etc.”). Many scenarios also took the form of tradeoff modelling early on: “What would be the consequence of not acting?”, highlighted in New Zealand and the UK, but of course paramount in every response.

#### Time series plots for past and future were paramount

Most modellers utilised time series plots with the left hand side displaying past and right hand side displaying future estimates, of for example bed occupancy.

#### A consistent fixed y-axis is important

In Australia, for example, consistent y-axes across the eight jurisdictions enabled comparison across jurisdictions. It was acknowledged that it is important to show scenario projections ‘fairly’, giving equal weight to different scenarios either by using a fixed y-axis across multiple plots or by overlaying on the same axes to facilitate comparison. One French modeller also at times would highlight the y-axis as this was more visually appealing and could drive a message home on high counts, for example, declining hospital bed capacity.

#### Central estimates and uncertainty need careful consideration

All respondents acknowledged that uncertainty was hardest to translate. However there were some novel suggestions from different teams. Many groups evolved to resist presenting a central estimate/ median or ‘line in the middle’, as this often caused overfocusing on exact values confusing the overall message. “We refused to show anyone a median! And what median do you choose anyway?”. One French team also remarked that they did not highlight any central estimate instead using lower or higher estimates to emphasise range over the point. New Zealand modellers similarly experimented with different ways of displaying uncertainty, and settled on presenting 10-15 trajectories, somewhat randomly chosen within a band. Displaying a sample plot of individual trajectories in this way worked well to communicate the concept *and* the possibility of variation.

This was stronger than a single curve or central estimate which would often interfere and be easily misinterpreted. Australian modellers had similar evolution in their visuals with discussion on quantiles vs. trajectories. They too overlaid a sample of trajectories, which after regular use, enabled policymakers to observe for example “you can see in the trajectories that there’s quite a lot of uncertainty this week”. Another important point was that confidence intervals can be more amenable to presenting as an aggregate numerical range, for example aggregate number of hospital admissions over six months as a numerical range accompanying the graph, rather than graphically.

However our study also identified a major counterexample to the aversion of best estimates. In South Korea, it was remarked that visualisations during the Delta wave showing multiple estimates were difficult for the government to interpret. “We are going up or going down?”. This led the researchers to choose to use best estimate curves in their presentations in the subsequent Omicron wave.

#### Thresholds and horizontal lines can be powerful

Some teams also evaluated the importance of horizontal lines on graphs to indicate certain thresholds. The most common was ICU bed capacity for hospitalisations plots. Horizontal lines were also used in for example Australian forecasts, using the BA1 ICU occupancy line to compare to the current BA4.5. This was supplemented with a table/ numerical outputs indicating the probability of exceeding the threshold over the time horizon of the forecast.

#### Off the range estimates may need explanation to maintain credibility

Curves exceeding the y-axis for example in times of exponential growth of cases (‘going off the graph’) were initially difficult to digest with policymakers in Canada and commentators suggested this undermined the credibility of the modelling. Over time, with available data, validation of the modelling increased confidence in modelling, and the concept of exponential growth was understood.

#### Consider greying to indicate lags

Teams in Canada and the UK used grey zones or bars to indicate lag in for example case reporting and confirmation.

#### Be consistent week to week

Lastly, interviewees agreed that consistency in colours, styles, graphs etc. is important. “Be consistent with the way you packaged the first information”. Presenting in the same format each week enabled policymakers and advisors to gain familiarity and to provide a pattern of feedback - “could you change this for next week?” and so on.

### Metrics

#### Decision makers need to know: Trends, Severity, Capacity

Heuristically, policymakers reported being concerned with three key factors: Trends, Severity, and Capacity. And for trends, what is the local vs global picture vs geographic spread? Timing is also really key - one wants to understand “when do we have to scale up our activities?” - a question that often pivots on hospital capacity in public health settings. Being able to give approximate lead times was appreciated, for example lead time for the peak in number of ICU beds.

#### Centre on familiar quantities; ‘doubling time’ and ‘time to X’ can be helpful metrics

Graphs should be centred around the numbers and quantities which the relevant decision makers are most familiar with. For example in the early COVID-19 response in France, this equated to presenting number of cases, number of hospitalisations, and number of deaths, as a function of time. In Uganda and the UK, although disparate countries in many ways, interviewees commented that doubling time and ‘time to X’ concepts (time for current number of cases to double, time to 100 cases, time to 1000 cases, etc.) were picked up much more smoothly by policy than the reproduction number ‘R’/’Rt’. It was remarked that this is because there is no extra knowledge needed to make use of these concepts. “Even for R, the greater than or less than one, that’s another fact I have to remember to make use of this fact”. “Rt estimation was interesting, but it was more relevant to be able to convert models to impacts on hospitalisations and deaths”. Streamlining advice by presenting modelling findings in terms of these more operational concepts (doubling time, time to X, hospitalisations, deaths) was preferred in a few different settings.

Key graphics in other COVID-19 settings included daily number of cases, positivity rate, and testing rate (Kenya); reproduction number against time in different cities, number of cases against time in different cities, and number of ICU beds (Colombia); and hospitalisations and deaths (New Zealand). For example Kenyan modellers showed visualisations to compare testing thresholds between WHO figures and Kenya, taken from the National Public Health Laboratory. This type of comparative visualisation was clearly understood by the Ministry of Health and mobilised resources for testing interventions. Number of cases and testing trends fed into the positivity rate, a simple graph which was heavily relied upon. Kenyan actors also made use of visualisations of key sectors such as hospitals to invoke action and interventions.

#### Counterfactuals as a novel retrospective exercise

A final suggestion was counterfactuals modelling, which was carried out by Canadian modellers after a suggestion from one of their ministers. Public Health action often aims to prevent and control, and it can be hard to explain what could have happened if a country had chosen not to act. Counterfactuals modelling was explored as a new way for supporting and promoting the utility of outbreak response work.

### Graph-reading and interpreting visuals

Policy and decision makers stressed that modelling can be used for *planning*. They stated it must be understood that visuals are “a simulation, not what will definitely happen”. Some policymakers commented that they have learnt a little already on how to visualise - for example “don’t take the values at face value”, “see as a trend”. Modelling provides a rough trend; one shouldn’t be concerned with exact numbers even though this is incongruous with policy at times. There was also a distinction made by modellers between scenario projections and forecasts - the outputs of these mean different things for policy and they should be read and interpreted separately. To combat the immediate response of number-focusing, some of the ‘translators’ and intermediaries in our study described their strategies for aiding their decision makers with interpreting visuals:

> “Trying to pull people away from just looking at where this curve is peaking”.
>
> “Look at the shape, look at the worst case scenario and the best case scenario, and therefore these are the things that matter”.
>
> “These are the patterns you can expect if this, this and this, THEN this will happen”.
>
> “A key variable or a key driver for this not happening will be this, whereas if this happens, then it will reinforce this pattern”.

Translators often focused on giving a *qualitative* result like the above quotes demonstrate; often text-led. It was commented that the actual policy translation is often qualitative and heuristic, such as “Will prevalence in region A be high, medium, or low?”.

### Examples

We conclude with some real-world examples of visualisations used in COVID-19 modelling. These are shown in Figures 2-5 below.

## 4 Discussion

This article provides a first evidence base for effective science communication to policy in crises. Results highlight crucial first steps to creating guidance on communication in epidemic response modelling, and many of the findings will also be widely applicable for other fields looking to introduce guidance for their own emergency response work. We have synthesised and presented advice on modelling visualisations - rapid actions that epidemiologists, scientists, and other knowledge brokers can implement to aid understanding *for policy* and ultimately improve the usability of outputs in crisis situations.

Key themes identified include the simplicity of visuals, something that almost all interviewees commented on without prompt, and the integrity of onward messaging or ‘re-translation’, ensuring visuals and presentations were robust to subsequent explanations through different levels of the policymaking process. We also observe a preference towards figures that would give the policymaker options, with numerous modelling teams presenting ‘scenarios’ or ‘options’ to their policymakers. Consistency in the style and format of visuals is encouraged week-to-week, and although interviews focused mainly on graphs and plots, many policymakers also commented that tables were another valuable method of visual communication.

In related literature, 95% of respondents from the aforementioned CMCC stakeholder survey group found graphical representations to be ’very helpful’ [8], matching the general consensus of our interviewees. The concept of simplicity was also at the forefront in CMCC findings; decision makers reportedly preferred “simple visual aids with summary messages”. One again sees discussion of appropriate metrics and a recommendation that modellers being able to produce ’top-line’ key-takeaway/ one-liner messages were helpful. The CMCC Policy report also suggests clear labelling for example of axes, avoiding double axes, and not crowding visuals. These ideas were confirmed by the policy and decision makers and science advisors of our own study. Additionally, heavily overlaid charts were rated lower by CMCC survey respondents for ‘ease of interpretation’ and ‘clarity’. Succinctly phrased in their report, “unless the chart is trying to display the relationship between the two variables, then creating two charts will be less prone to misinterpretation”.

Looking further back, in Australia efforts to improve communication have been longstanding and communication has been developed over 15 years. Interviewees spoke of dedicated research prior to the pandemic to understand what was most useful for policy makers, in partnership with the Melbourne School of Government [20]. For example, long reports were deemed necessary for security and confidence in the Australian setting but impractical for time management. There was also further work on visual preferences, identifying which graphics were appeasing and easier for the local policymakers to understand. We advocate for further dedicated research projects on visualisation and science communication to different groups of policymakers and decision stakeholders.

Our own study is not without limitations. We note that the terms ‘policy’ and ‘government’ are used loosely as due to the limited scope of our study, we were unable to stratify results by the different levels of policy actors that make up a government. Results instead represent only general advice for presenting epidemiological results to any kind of policy actor.

We also acknowledge that results in this article are primarily directed towards policy spaces where there is low to moderate ‘modelling literacy’ (i.e. where policy and decision makers have a limited understanding of the underlying methods of infectious disease modelling, due to it not being their expertise or remit). However, our study observed a range of modelling literacy in the different government groups that were examined, including a rare setting where policy and decision makers had extensive training and familiarity with both epidemiological concepts and the mathematical models themselves.

We note that state-level modelling, multi-country modelling, and settings where modelling did not contribute to *national* COVID-19 response were all outside the scope of this study. Multi-country modelling efforts such as that of the CoMo Consortium [21] and IHME [21,22] may require presentation to multiple stakeholders with differing cultural and political climates - nonetheless the high-level results presented in our study are likely to be applicable at multiple scales.

In terms of methodological limitations, qualitative research is not definitive or exact and one should acknowledge the researcher’s role in collecting, interpreting, and analysing the data. In line with good practice in qualitative interviewing [23], we also acknowledge the interviewer’s developing interview style and interviewing technique throughout data collection, which ran from May 2023 to March 2024. A number of measures were taken to ensure academic rigour: audio recordings were used to enable transcription and accurate note-taking, all transcripts were coded and independently checked by members of the research team, quotes used in the final analysis were compared back with the original transcripts to ensure meaning had been maintained, and all interviewees were invited to fact-check the final analyses.

Notwithstanding these limitations, this study serves as a first evidence base for presenting scientific and modelling visualisations. This should now be strengthened with the current best knowledge on Visual Processes and further dedicated research to substantiate key actors’ advice. This study will directly contribute evidence to formal activities developing best practice on the visualisation and translation of modelling, and it is hoped that the article also provides evidence on which adjacent scientific fields can develop their own policy communication guidelines.

## Author contributions

LH proposed the study. LH, AT, OR, and SF were involved in conception and design. LH carried out the data collection. LH, CR, and AT performed the analysis. LH wrote the first draft of the manuscript. All authors provided feedback on the manuscript and approved the final version for publication.

## Acknowledgements

LH acknowledges support from the Wellcome Trust (block grant no. RG92770). SF also acknowledges support from the Wellcome Trust (grant no. 210758/Z/18/Z). The authors also thank Sungmok Jung, Mircea Sofonea, and all other interviewees for dedicating their time and experience to the study.

The authors declare no conflicts of interest.

## Data availability

The available anonymised data has been presented in this manuscript and any associated manuscripts where possible.

## Footnotes

1 For brevity, we use the term ‘country’ to mean ‘country or jurisdiction’ throughout this article, with the knowledge that Hong Kong, China is a jurisdiction or ‘special administrative region’ and not itself an independent country.

## Notes

### Competing Interest Statement

The authors have declared no competing interest.

### Author Declarations

This research study has been reviewed by the University of Cambridge Psychology Research Ethics Committee (application number PRE.2023.034).

### Summary of Updates

Minor edits to style to prepare for journal submission.

