## Supplementary Material for "Visual preferences for communicating modelling: a global analysis of COVID-19 policy and decision makers"

**Interview themes and questions**

**Study title**: “The use of infectious disease modelling in Covid-19 response: a multi-country study”

The themes and questions below will be used as prompts to guide our interview discussion. Opening questions are in blue, example follow-on questions in orange. There will also be ample opportunity to discuss any other information that you feel is relevant, helping us to understand the interaction of modelling and policy during Covid-19 in your country of practice.

***Structures and pathways to policy***
*How did the collaboration with modelling begin? Is this the first time infectious disease modelling has been used to advise the government in your country?*

- *Do you feel that the setup was effective? Was it easy for modelling results to reach senior government? And could policymakers easily suggest questions of interest for modelling?*

***Communication and visualisation***
*What level of communication occurred between yourself and senior government, and yourself and modellers throughout the pandemic? For example, did you have the opportunity to discuss/report model findings with senior government on a regular basis and to feedback to the modelling teams?*

- *Translation. Who took up the role of translation (converting complex epidemiological findings into meaningful language for decision makers)?*
- *Visualisation. What aspects of graphs and plots were most helpful to decision-makers and advisors?*
- *Can you give an example of a plot that you felt worked well? And an example of one that was perhaps misunderstood by policymakers or not quite so effective at demonstrating model findings?*
- *Selective publishing. How do you decide what to include or exclude in reports/presentations to government? Level of detail? How accurate do you choose to be?*

***Collaboration and knowledge transfer***
*Do you feel you had access to sufficient country expertise for infectious disease modelling?*

- *Cross-disciplinarity. Were modellers encouraged to work closely with other scientists informing the response or were the different streams of evidence kept deliberately distinct? Economics/ behavioural science/ medicine etc.*

***Evaluation and reflection***
*What are your key takeaways or lessons learned from working in Covid-19 and supporting modelling? What was your biggest mistake (in relation to modelling)?*

- *Can you give an example of where you feel modelling got it wrong?*
- *Has there been interest in reflecting on how modelling can be made more useful to public health decision-makers? Perhaps through improved pathways, improved translation, and so on.*
- *Future of modelling. Do you think modelling will be used in future outbreaks in your country? What advice would you give to future actors?*
